# Adapting cleft care protocols in low- and middle-income countries during and after COVID-19: a process-driven review with recommendations

**DOI:** 10.1101/2021.10.14.21265004

**Authors:** Matthew Fell, Michael Goldwasser, B.S Jayanth, Rui Manuel Rodrigues Pereira, Christian Tshisuz Nawej, Rachel Winer, Neeti Daftari, Hugh Brewster, Karen Goldschmied

**Affiliations:** CLEFT Charity, Chelmsford, United Kingdom; Cleft Collective, University of Bristol, Bristol, United Kingdom; Operation Smile, Virginia Beach, USA; Craniofacial and Surgical Care, University of North Carolina School of Dentistry, Chapel Hill, NC, USA; ABMSS, Bengaluru, India; Faculdade de Medicina da Universidade de Sao Paulo, Sau Paulo, Brazil; Instituto de Medicina Integral Prof Fernando Figueira, Recife, Brazil; Cliniques Universitaires de Lubumbashi, Democratic Republic of Congo; Transforming Faces, Toronto, Canada; Hospital Dr Luis Calvo Mackenna, Santiago de Chile, Chile; Project Harar Ethiopia, Henfield, United Kingdom; Cleft Lip and Palate Program, Yekatit 12 Hospital Medical College, Addis Ababa, Ethiopia; CORSU Rehabilitation Hospital, Kisubi, Uganda; Smile Train Global Medical Advisory Board, New York, USA; Division of Plastic Surgery, Michael E. DeBakey Department of Surgery, Baylor College of Medicine, Department of Surgery, Texas Children’s Hospital, Houston, Texas, USA; Noordhoff Craniofacial Foundation, Taipei, Taiwan; Craniofacial Center, Plastic and Reconstructive Surgery, Chang Gung Memorial Hospital, Taoyuan, Taiwan; Great Ormond Street Hospital for Children, London, United Kingdom; Division of Plastic and Reconstructive Surgery, The Hospital for Sick Children, Toronto Canada; The University of Toronto, Toronto, Canada

**Author notes:** **Corresponding author:** Matthew Fell, The Cleft Collective, Bristol Dental School, University of Bristol, Oakfield House, Oakfield Grove Bristol, BS8 2BN, United Kingdom.

**Keywords:** comprehensive cleft care, low- and middle-income countries, COVID-19, Circle of Cleft Professionals

## Abstract

**Objective:** A consortium of global cleft professionals, predominantly from low- and middle-income countries, identified adaptions to cleft care protocols during and after COVID as a priority learning area of need.

**Design:** A multidisciplinary international working group met on a videoconferencing platform in a multi-staged process to make consensus recommendations for adaptions to cleft protocols within resource-constrained settings. Feedback was sought from a roundtable discussion forum and global organisations involved in comprehensive cleft care.

**Results:** Foundational principles were agreed to enable recommendations to be globally relevant and two areas of focus within the specified topic were identified. First the safety aspects of cleft surgery protocols were scrutinised and COVID adaptions, specifically in the pre and peri-operative periods, were highlighted. Second, surgical operations and access to services were prioritized according to their relationship to functional outcomes and time-sensitivity. The operations assigned the highest priority were emergent interventions for breathing and nutritional requirements and primary palatoplasty. The cleft services assigned the highest priority were new-born assessments, paediatric support for children with syndromes, management of acute dental or auditory infections and speech pathology intervention.

**Conclusions:** A collaborative, interdisciplinary and international working group delivered consensus recommendations to assist with the provision of cleft care in low- and middle-income countries. At a time of global cleft care delays due to COVID-19, a united approach amongst global cleft care providers will be advantageous to advocate for children born with cleft lip and palate in resource-constrained settings.

## INTRODUCTION

Cleft lip and/or palate (CL/P) is the most common craniofacial congenital anomaly, occurring in approximately 1/700 live births worldwide (Mossey et al., 2009). If untreated, CL/P is highly problematic for children and their families as it gives rise to functional difficulties with speech, eating, social interaction and child development. It is well established that the best way to treat a child born with CL/P is a multidisciplinary team (MDT) of specialised professionals following a protocol of comprehensive cleft care (Kassam et al., 2020). Unfortunately, global inequalities exist, with provision and access to comprehensive cleft care differing depending on geographical location of birth (Sharratt et al., 2020). Low- and middle-income countries (LMICs) face unique challenges due to the existence of constrained resources (Ma et al., 2020) and data collected in 2014 estimated the backlog of untreated cleft in LMICs to be more than 600,000 cases (Carlson et al., 2016).

On March 11^th^ 2020 the World Health Organisation declared COVID-19 to be a global pandemic. This had a major impact on healthcare systems and services were accordingly re-prioritised, with emergency and trauma services continuing but many elective procedures being delayed or postponed (American Cleft Palate-Craniofacial Association, 2020a, 2020b; Cleft Development Group, 2020). The pandemic has undoubtedly exacerbated the backlog of healthcare interventions for children born with CL/P, as they are for the most part regarded as planned elective procedures, although the magnitude of these delays on a global scale is yet to be fully appreciated (Stoehr et al., 2021). Projections using data from 67 LMICs estimated 25,000 fewer cleft operations performed during 2020 compared to 2019 (Vander Burg et al., 2021). In Peru, children born with CL/P were having primary reconstructions at a significantly older age during the pandemic when compared to a pre-pandemic cohort, with delays most marked in primary cleft lip and nose reconstruction (Rossell-Perry & Gavino-Gutierrez, 2021). Prioritising cleft care in the overcrowded healthcare system when the pandemic ends will be challenging, even in high resource settings (Breugem et al., 2020). LMICs are likely to face additional barriers to reinstating elective cleft services, which may include access to COVID testing, treatment, vaccines, personal protective equipment (PPE) and travel restrictions impacting most upon patients living in remote rural locations (Ramanathan et al., 2021; Stoehr et al., 2021).

The Circle of Cleft Professionals (CoCP) is a coalition of international non-governmental organisations (NGOs), which aims to support healthcare workers around the globe to provide comprehensive cleft care (Circle of Cleft Professionals, 2021a). On September 17^th^ 2020, CoCP facilitated an international virtual conference entitled ‘Solutions for Comprehensive Cleft Care: Responding to COVID’. Following the conference, an online CoCP COVID-19 Survey was designed, aiming to identify challenges that cleft professionals face in light of the pandemic, particularly in LMICs, and to identify learning priorities (see supplementary material for copy of survey). The survey was translated into 6 different languages to facilitate broad representation and disseminated internationally online in February 2021 to global cleft professionals through a network alliance of 10 global NGOs. The survey received 175 responses, 74% of which were from cleft professionals located in one of 40 LMICs (see Supplementary Figure 1). A priority area identified for further learning from the survey was ‘adapting COVID-19 cleft care protocols in light of evidence-based research’.

A clinical protocol (also known as a plan, pathway or guideline) is a tool to guide evidence-based healthcare (Rotter et al., 2019). A protocol aims to standardise care and has the potential to streamline multidisciplinary clinical practice by detailing steps of management. CL/P is associated with a striking diversity of management protocols in common use and furthermore there is a paucity of a scientific evidence to support any of them (de Ladeira and Alonso, 2012; Hardwicke et al., 2017). The reason for this may be the complex, heterogeneous nature of the condition, with multidisciplinary care administered by a range of specialists at different stages of child development (Allori et al., 2017). There are examples of individual cleft centres, such as in Adelaide and Lima, publishing their protocols (Rossell-Perry and Luque-Tipula, 2020; Schnitt et al., 2004) and also nationwide cleft standards, which detail threshold age targets for the completion of primary operations (NHS England, 2018). It is perhaps not surprising that consensus for international standardisation has not been reached for the delivery of cleft care protocols, nor for the assessment of outcomes (Weidler et al., 2021). The World Cleft Coalition, formed from several international NGOs, has published international treatment program standards with a primary focus on the delivery of ethical, safe, accessible and patient-centred care (Kassam et al., 2020). The coalition purposefully did not dwell on protocol technique and timings, due to the well documented controversies in this area, but instead attempted to make balanced recommendations to allow for the levels of resources available locally.

The need to adapt aspects of the cleft protocol during and following COVID-19 has been identified by global partners and is important in the quest towards re-establishing international comprehensive cleft care services. The CoCP platform was used to bring together cleft professionals from diverse locations to consider adaptions to the cleft care protocol by pooling experience and reviewing available evidence. The overall aim was to formulate practical consensus recommendations to help providers in LMICs to deliver comprehensive cleft care protocols during and after COVID-19.

## METHODS

### Process overview

A multi-stage process, designed specifically for this context by CoCP organisers and advisors, is summarised in a flow diagram in Figure 1. The process was centred around the formation of working groups to consider topics highlighted in the CoCP COVID-19 Survey. The application to participate in a working group was disseminated widely through the CoCP membership and beyond. Applicants were placed in one of six working groups based on research interests, fluency in English or Spanish, and in an attempt to ensure diversity of professional context, discipline, geography and NGO affiliation. Working group members were orientated into the process and encouraged to consider their allocated topic area before meeting collectively on three separate occasions over a six-week period in 2021. The process culminated with a presentation of recommendations at a round table within an international virtual conference, that had free registration and was widely advertised, entitled ‘Solutions for Comprehensive Cleft Care: Covid and Beyond’ on June 2^nd^, 2021 (Circle of Cleft Professionals, 2021b).

**FIGURE 1.**
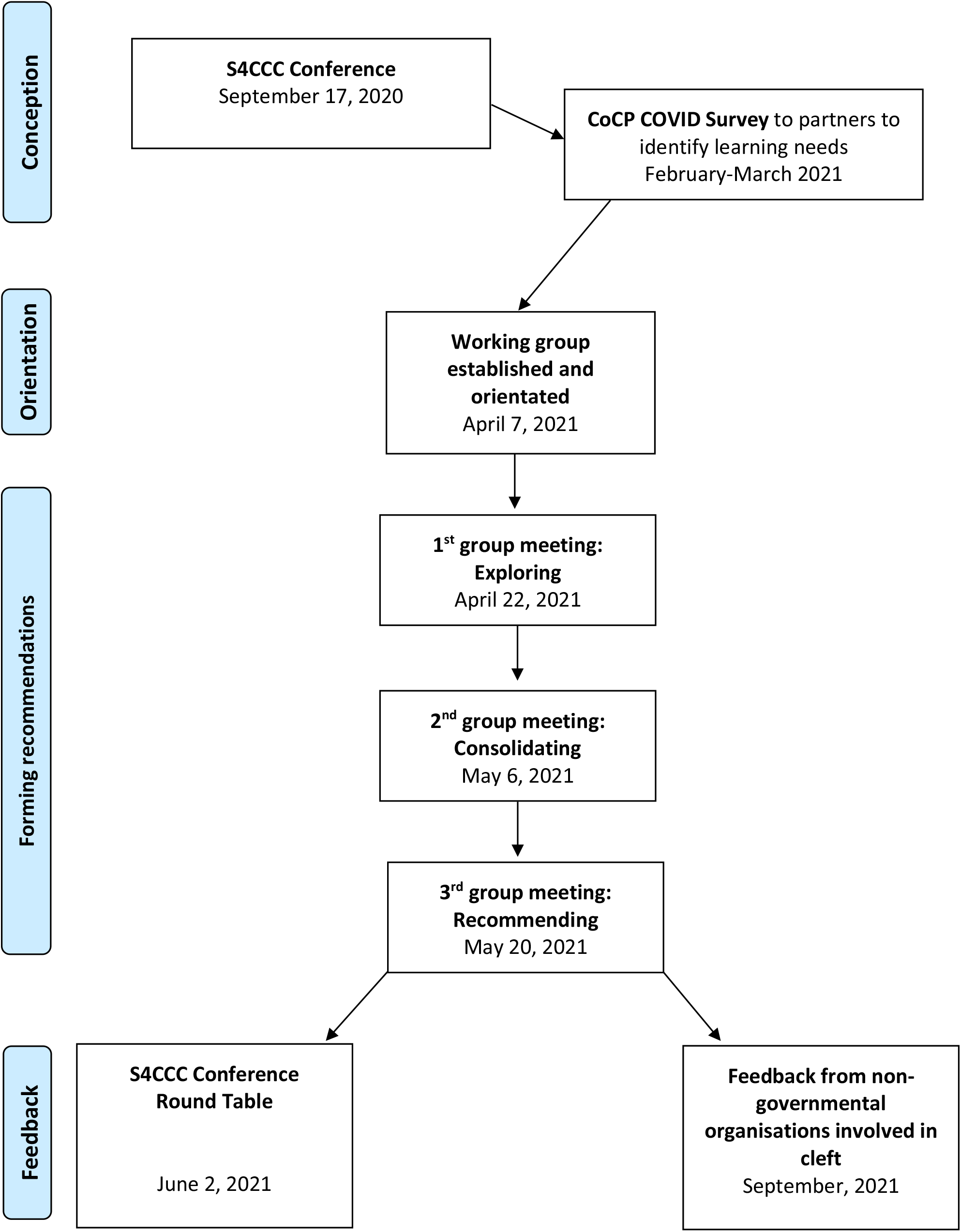
A flow diagram to describe an overview of the process used

The structure of this report was inspired by the World Cleft Coalition publication (Kassam et al., 2020) as it was considered a rare example of an international collaborative endeavour in global cleft care and the benefit of a similar format for end user interpretation and application was recognised.

### Composition of the Working Group

This working group was composed of seven individuals; six healthcare professionals and one non-healthcare professional in an administrative role (see Table 1). There was representation from seven countries in four continents and inclusion of three speciality areas from the cleft multidisciplinary team. Working group members had a range of experience in the delivery of comprehensive cleft care within their own countries and overseas and were affiliated with a range of global cleft organisations.

**Table 1:**
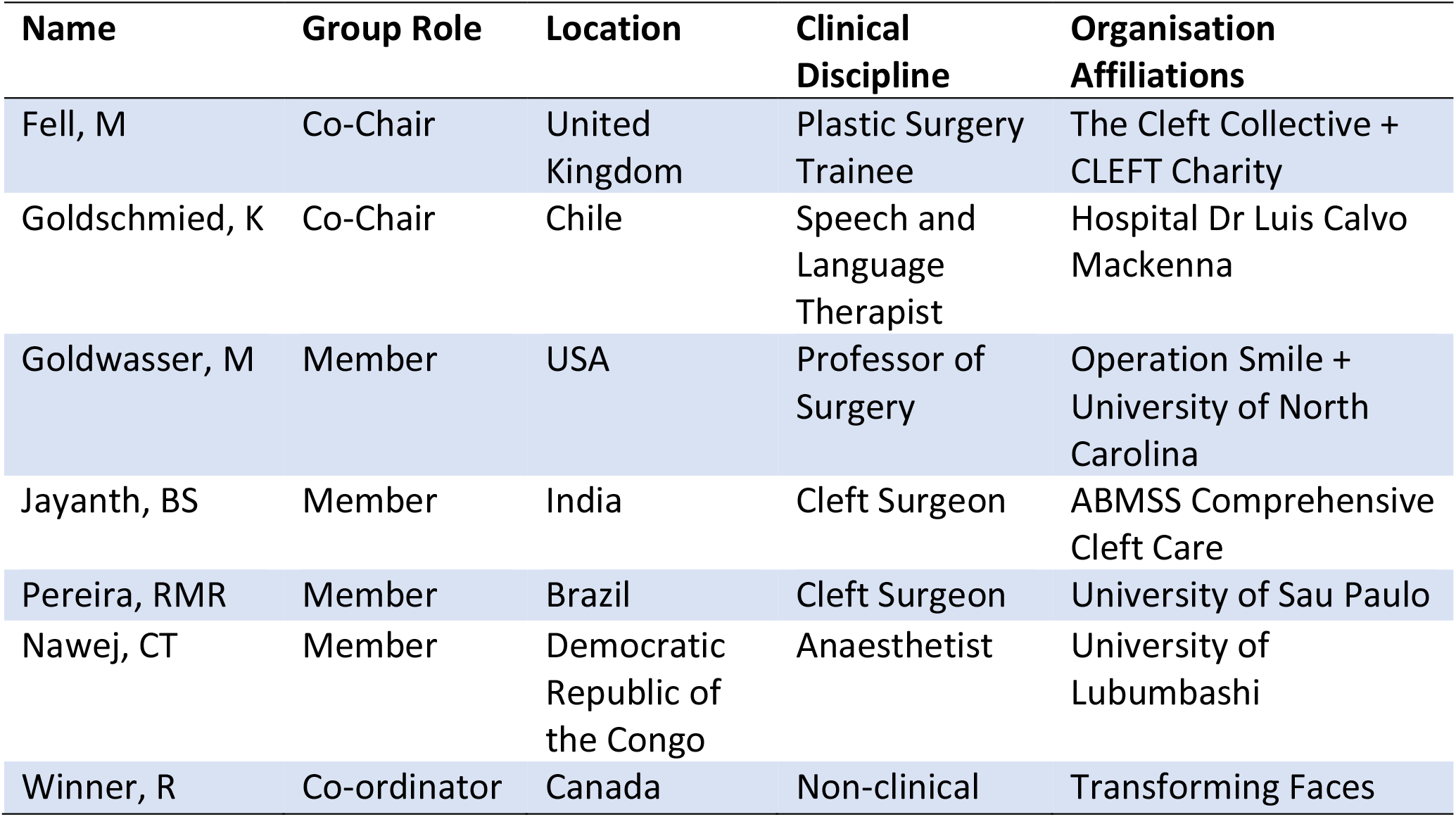
The composition of the working group

### Making and testing recommendations

The working group met virtually on three occasions using a videoconferencing platform. The first session entitled ‘exploring’ involved open discussion of the assigned topic and highlighting areas within the assigned topic in which to focus. The action plan from the first meeting was to identify available guidance through literature searches in combination with personal experience and organisational contacts. Literature was categorised according to levels of evidence (see supplementary Figure 2) and shared between group members in the interim period to stimulate discussion via email, instant messaging and an online conference platform. The second meeting entitled ‘consolidating’ involved consideration of the identified evidence and the creation of preliminary consensus recommendations. The final meeting entitled ‘Recommending’ consisted of reviewing and refining the group consensus recommendations. At the culmination of this process, the working group presented their recommendations at conference round table and attendees were encouraged to comment and provide feedback. The round table enabled a pilot test of the recommendations and an opportunity for feedback from the attending audience. Further feedback was sought from leading cleft professionals allied to the CoCP NGO network. The feedback was used to help understand the global implications of the recommendations and refine them as needed.

## RESULTS

The working group considered the topic area of ‘adapting COVID-19 cleft care protocols in light of evidence-based research’. A consensus was reached on foundational principles and recommendations made in two focus areas: first, surgical safety measures within the cleft care protocol and second, prioritisation of surgical procedures and access to cleft care services.

### Foundational Principles

The working group agreed that recommendations for cleft protocol adaptations, supported by a body of identified scientific evidence, could be beneficial to help coordinate and unify the international lobbying of policy makers regarding the need for comprehensive cleft care provision during and after the COVID-19 pandemic in LMICs. The target audience were global cleft care providers in resource-constrained settings, which includes health care professionals and/or management teams at a regional, national or international cleft service delivery level. The aim was to create a document that would be a helpful aid to global lobbying efforts in LMICs, with an appreciation that recommendations could neither be comprehensive nor specific to reflect the needs of each individual healthcare system and setting.

Potential pitfalls were recognised with recommendations relating to global cleft care protocols. First, it was clear that cleft protocols would vary enormously in resource-constrained settings, with influencing factors including the setup of local healthcare services, the socio-economic context, the availability of multidisciplinary care and dependence on external teams for the provision of cleft care. Each nation has its own government, healthcare laws and potential existence of additional crises, such as civil war, which would have a significant impact on the delivery of any healthcare protocol. There was an endeavour to make protocol recommendations that would be broadly applicable, non-judgemental and evidence-based by referring to relevant literature and guidance. Second, the contentious nature of many aspects of the cleft care protocol was acknowledged, especially with regard to timings, sequences and techniques in use. Prescriptive statements were avoided, with recommendations made instead according to widely accepted principles. The hope was that the recommendations would facilitate the provision of cleft care during and after COVID-19 in LMICs, rather than adding restrictive measures for healthcare providers.

The working group focused upon two areas within the cleft care protocol that were felt to warrant the greatest need; the first focus area was operative safety, and the second was prioritisation of surgical procedures and MDT cleft services.

### Focus area 1: Surgical safety adaptions to the cleft care protocol

The primary focus of any healthcare protocol is to promote the safety of patients, their family and the healthcare providers. Following the inevitable delays in cleft care provision following COVID-19, the reinstatement of cleft services must be done safely and according to the latest available evidence. Many aspects of safety were in place before the pandemic and for the most part, these would continue during or after the pandemic with some notable additions and considerations. Recommendations centred upon suggested additional adaptations to be considered during and after COVID-19 and have been categorised according to the period of operative care (pre, peri and post) as described in Table 2.

**Table 2:**
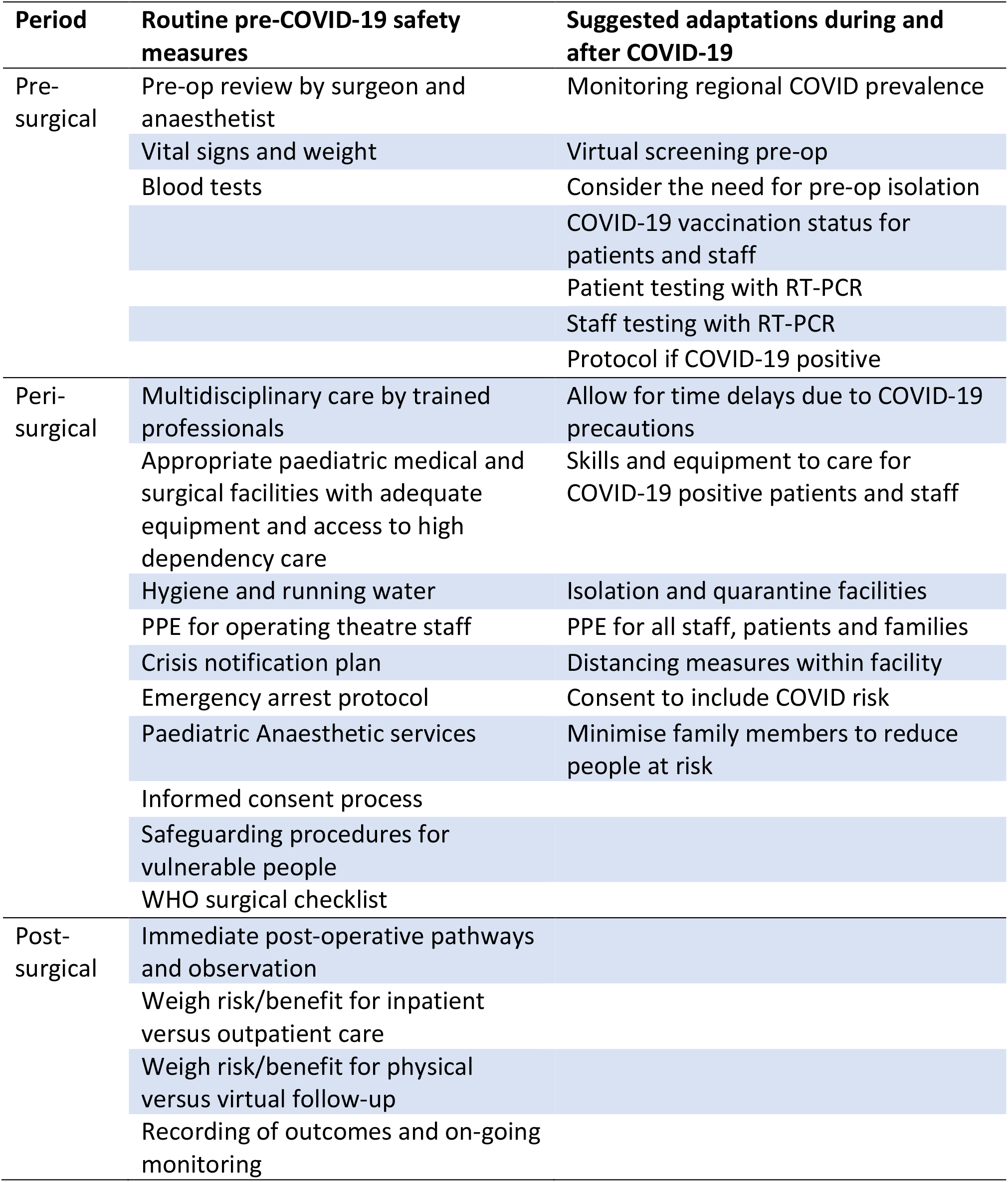
Cleft surgery safety measures that were routine before COVID-19 and specific adaptations for consideration during and after COVID-19

**Table 3:**
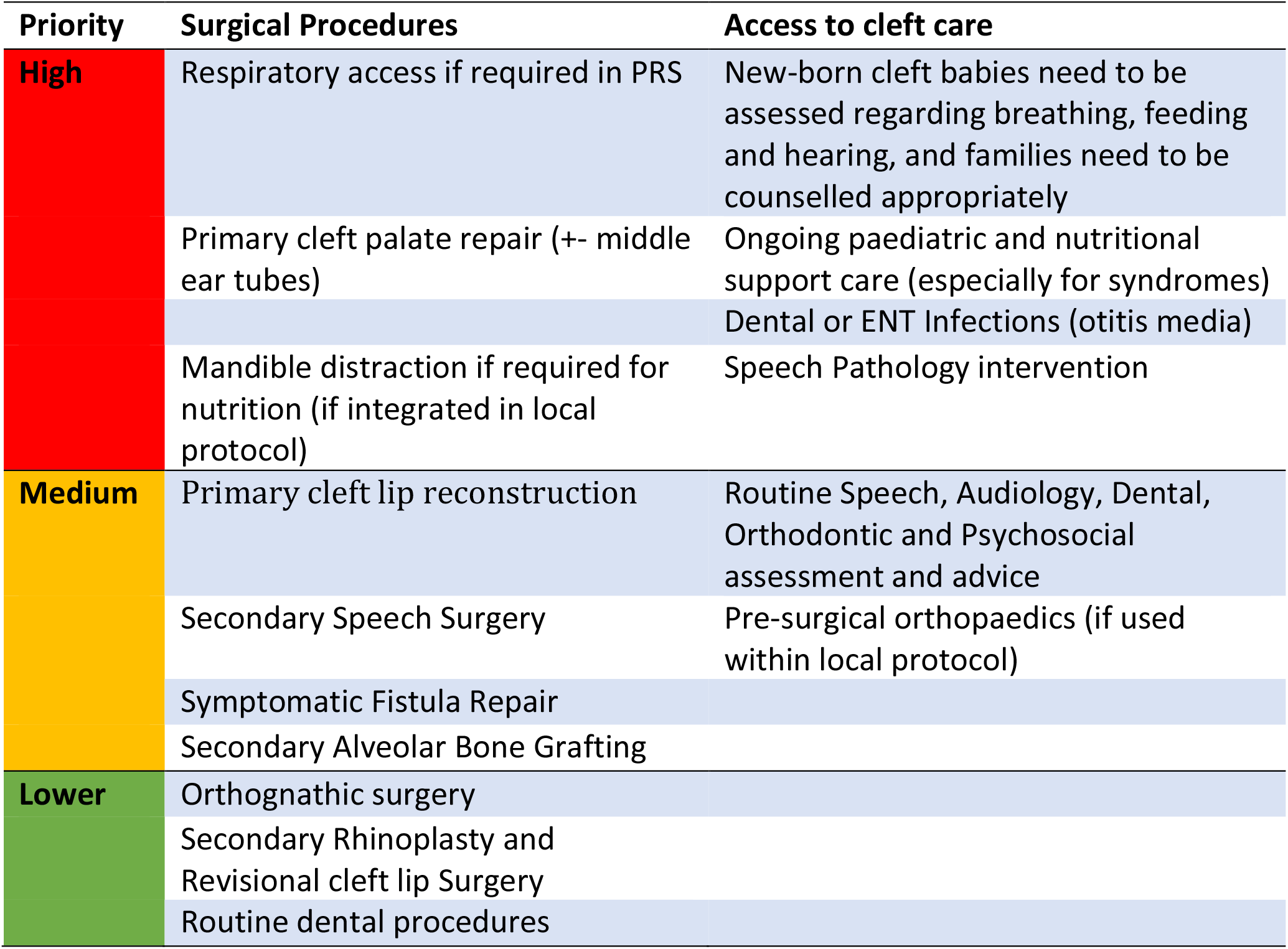
Recommended time-sensitive prioritisation of surgical procedures and access to comprehensive cleft care

Pre-operative safety protocols exist to assess whether the patient is safe to proceed with a procedure and often incorporate a consultation and basic tests. Post-COVID-19, pre-operative assessments need to be expanded to judge the risk of the virus causing harm to patients, families and providers. The extent of pre-operative modifications (such as frequency of COVID-19 testing and the need for isolation strategies) can be adapted in response to regional COVID-19 prevalence, which has been classified as low (<0.5%), medium (0.5-2%) and high (>2%)(Royal College of Paediatrics and Child Health, 2020). Whilst an in-person consultation with the patient, surgeon and anaesthetist remains vital, virtual screening for COVID-19 symptoms can be successfully utilised (Royal College of Paediatrics and Child Health, 2020). COVID-19 testing, performed as close to the time of care as possible, is an important adaptation of the pre-care protocol, whilst recognising the need for flexibility due to access to testing facilities. Establishing vaccination status is important but vaccine availability will likely be a challenge in LMICs due to global inequity and therefore an emphasis on PPE for patient, families and staff may be required (Ma et al., 2020; Stoehr et al., 2021).

Peri-operative safety protocols exist to maintain patient well-being whilst under the care of health professionals. Healthcare systems are well accustomed to protocols relating to safety during this period and should have training and equipment in place to deal with adverse events(Operation Smile, 2020; Smile Train, 2018.). The World Health Organisation (WHO) has published guidance on equipment and facilities required to run a safe surgical service (World Health Organisation, 2003). Securing adequate stocks of PPE is an important element of creating a safe working environment but will likely be a challenge amongst other resource shortages in LMICs (Ma et al., 2020). Adaptions are required to factor in the space, facilities and time to address COVID-19 risk reducing precautions such as social distancing and isolation (Cai et al., 2021). Specifically, consent for procedures should detail the risk of contracting COVID-19 during the hospital stay and emphasise the importance of following current COVID-19 guidance (Ramanathan et al., 2021).

Post-operative safety protocols exist to ensure that the surgical care episode was successful and that the patient does not develop complications that require treatment. The decision to follow-up patients in person or remotely is made on the merits and practicalities of both options and has many influencing factors, of which COVID-19 is just one. Irrespective of COVID-19, it remains important that operative outcomes are accurately assessed and recorded and indeed the advances in telemedicine during the COVID-19 pandemic may ultimately make this easier. Arguably, there may not be any specific safety adaptions required in this post-operative phase of the protocol during or following COVID-19 but maintaining levels of follow-up surveillance when resources are restricted may be a challenge.

### Focus area 2: Prioritisation of surgical procedures and MDT cleft services

There is a need for prioritisation within the cleft protocol despite each element of comprehensive cleft care having equal importance because some elements are time-sensitive and linked to functional outcomes, therefore their delay would lead to irreversible harm(Rossell-Perry and Gavino-Gutierrez, 2021). Prioritisation of care according to clinical urgency has been widely encouraged as a vital part of re-establishing elective services amidst the backlog of untreated cases (Royal College of Paediatrics and Child Health, 2020). Elements of the cleft protocol were prioritised primarily based on time-sensitive functional outcomes, whilst also recognising the importance of aesthetic and psychosocial outcomes.

#### Prioritisation of surgical procedures

Surgical emergencies for patients born with CL/P, such as airway or nutritional compromise, require potentially life-saving surgical interventions and are therefore an obvious priority. The airway can be compromised in Pierre Robin Sequence, primarily due to glossoptosis and emergent surgical procedures to secure the airway, although rare, may be required (Breugem et al., 2016). The utilisation of mandibular distraction osteogenesis for children with micrognathia to improve breathing and eating is more controversial, with long-term outcomes in facial development yet to be determined (Breik et al., 2016), but was prioritised because of it aims to improve vital functions, with the proviso that it formed a part of the agreed local protocol (Ramanathan et al., 2021).

Primary palatoplasty was considered a high priority due to the body of literature identified to demonstrate its relationship with both speech and maxillary growth outcomes (see supplementary table 1 and 2). Evidence suggests the palate needs to be functional when sounds are first learned in order to avoid the development of compensatory speech patterns (Chapman et al., 2008). The optimal primary palatoplasty regime is a source of continued debate (Lohmander et al., 2012; Rohrich and Byrd, 1990) and randomised control trials currently in process aim to define the optimal timing for palatoplasty (Conroy et al., 2021). The SCANDCLEFT trials found that both good and poor functional outcomes can be achieved by a variety of palatoplasty techniques, sequence and timings and concluded that it was probably the operator skill and familiarity with the protocol that was most important (Shaw & Semb, 2017). Therefore, primary palatoplasty should be performed as a priority according to the accepted techniques, and within the scheduled timeframe, of the local cleft care protocol.

Primary cleft lip repair was categorised as a medium priority as earlier lip repairs have been shown to benefit mother-infant interactions and bonding (Murray et al., 2007). Secondary speech surgery, symptomatic fistulae repair and secondary alveolar bone grafting were medium priorities due to their time-sensitive association with functional outcomes of speech and maxillary growth, although they occur at an older age and with a wider window of opportunity when compared to primary palatoplasty (Breugem et al., 2020). Secondary speech surgery and the repair of symptomatic fistulae may be warranted before the child enters primary education with an aim to achieve normal speech to help optimise educational performance (Sell et al., 2015). Secondary alveolar bone grafting is commonly timed according to the decent of the deciduous canine tooth at approximately 8-12 years of age and aids the functional development of the alveolar arch to provide support for facial structures (Semb, 2012).

Orthognathic surgery, secondary rhinoplasty, revisional lip procedures and routine dental procedures were categorised as a lower priority, not to undermine their importance, but because they are not as acutely time sensitive.

#### Prioritisation of access to cleft care services

New-born babies with CL/P need to be assessed regarding breathing, feeding and hearing and this is a priority, both for the health of the baby and to provide support for parents during this critical neonatal period. Some children with CL/P, especially those with syndromes, will require ongoing input from medical professionals with paediatric experience. Acute dental infections or otitis media were prioritised because efficient treatment reduces the likelihood of permanent damage to dentition and hearing(Kuo et al., 2013). Speech pathology intervention was categorised in the highest priority to reflect the importance of speech outcomes and evidence to suggest that speech interventions reduce speech errors commonly observed in children with cleft (Sell et al., 2017). Innovations in telemedicine during COVID-19 have shown promising signs of the efficacy of delivering speech therapy remotely and this may be a great opportunity in LMICs going forward, especially for patients living in remote rural locations (Camden and Silva, 2021; Law et al., 2021; Pamplona and Ysunza, 2020).

Routine MDT assessments in dentistry, audiology, orthodontics, speech, psychology and surgery, as available within the local cleft team, were categorised as a medium priority because of the ability of these services to be delivered over a greater timescale without compromising outcomes. Pre-surgical orthopaedics was also categorised as a medium priority because despite its aim to improve tissue position and ultimately functional outcomes, it is not utilised universally, partly due to availability and partly due to the controversies surrounding efficacy (Hathaway and Long, 2014).

## DISCUSSION

### Overview of process

The structured process used in this study provided a positive collaborative experience, which should be encouraged in future global cleft endeavours. The condensed 6-week time period, with a pre-established ‘finish-line’, and a platform for the working group to present its recommendations, helped to increase intensity and provide urgency to the process. It became apparent that the variety of experience in the management of both CL/P and COVID-19 provided a rich environment for discussion and mutual learning. Scheduling meetings on the videoconferencing platform at the same time and day of the week helped to provide consistency and improve attendance, given the working group members’ multiple time zones and working commitments. It was helpful to specify focused aims from the outset of the process and to set tangible action points at the end of each group meeting. Encouraging continued discussion and the sharing of resources on virtual platforms between meetings helped to increase productivity. Consensus was achieved via identifying global areas of commonality and recognising areas of diversity and controversy.

### Summary of recommendations

The working group was tasked to make recommendations regarding the adaptation of cleft care protocols during and after COVID-19 to help facilitate the provision of global comprehensive cleft care in LMICs. Foundational principles were set to respect the complex multidisciplinary nature of cleft care in resource-constrained settings and the specifics of local protocols, as it has been demonstrated that familiarity with a protocol is of primary importance for the achievement of good outcomes (Shaw and Semb, 2017). Within the broad topic of cleft protocols, the two areas that were focused upon were safety and prioritisation. First, recommendations about adaptations to surgical safety protocols were made that were categorised into pre, peri and post-operative phases. Adaptations are most likely to be required in the pre and peri-operative phases to identify and manage COVID-19 risk. Second, recommendations to prioritise surgical procedures and access to cleft services based on time-sensitivity and functional outcomes. Primary palatoplasty was prioritised due its intimate relationship with speech and maxillary growth outcome. Infant medical services, management of acute infections and speech pathology interventions were the most highly prioritised cleft services.

### Interpretation and implications

The WHO has documented the far-reaching impact of the COVID-19 pandemic in terms of the widespread disruption to essential health services, but elective services are being re-established (World Health Organisation, 2021a). Global providers of cleft care will have to be prepared to adapt protocols to enable the comprehensive delivery of this essential health service. The literature and data on CL/P and COVID-19 is unsurprisingly sparse, given the relative infancy of the pandemic. (Salehi et al., 2021) have published recommendations for cleft and craniofacial outreach programs during the COVID-19 era with considerations for visiting teams before, during and after their visit away from their home country. The recommendations in current study focus instead on two important areas of the cleft protocol, and whilst applicable to visiting teams, are aimed at a wider audience of global cleft care providers in LMICs.

Safety is recognised to be of utmost importance when delivering cleft care (Kassam et al., 2020). The COVID-19 pandemic presents a safety dilemma because of the need to minimise the risk of the virus whilst balancing the risk of cleft treatment delays. The WHO has developed a useful facility assessment tool to enable rapid assessment of healthcare facilities to aid the provision of essential health services during the COVID pandemic (World Health Organisation, 2021b). More specifically for cleft, (Cai et al., 2021) reported management strategies to minimise the spread of the coronavirus during CLP treatment episodes in Shanghai. The collaborative group looked at safety protocols in common use before the pandemic and made recommendations on adaptations to consider specifically for COVID-19. Some of these adaptations, such as COVID-19 testing, will come at an increased monetary cost, and this is likely to be problematic in LMICs, where resources were already limited (Rossell-Perry and Gavino-Gutierrez, 2021). On the other hand, some COVID-19 adaptations represent innovations and the advances in telemedicine in particular, which has proven to be successful for pre-operative COVID-19 screening and speech therapy delivery, may be well suited to LMICs (Ramanathan et al., 2021). Vaccinations provide a crucial part of the international COVID-19 response, and the current global vaccination inequity will stand to reduce access to comprehensive cleft care for children born in LMICs (Circle of Cleft Professionals, 2021c).

In a crowded healthcare system following delays to many areas of planned services, prioritisation of care will be vital. Breugem et al., (2020) conducted a survey of cleft priorities during COVID-19 with 218 cleft professionals in Europe, Asia and the USA. The respondents viewed airway intervention for Pierre Robin Sequence to be an emergency procedure. Primary palatoplasty was similarly thought to be a priority, but there was no consensus about timing, with 70% recommending before 15 months of age and 22% before 18 months of age. Speech surgery, alveolar bone grafting, placement of ear tubes and primary cleft lip repair were viewed to be time dependent and therefore warranted prioritisation. In the United Kingdom, all surgical procedures were prioritised into four categories of urgency by the Federation of Surgical Specialty Associations in July 2020 to expediate the recovery of surgical services during COVID (Federation of Surgical Specialty Associations, 2021). Primary palatoplasty and secondary speech surgery were initially categorised as priority 3 but were upgraded to priority 2 in February 2021 (see supplementary table 3) following advice from UK cleft professionals regarding the association with functional speech outcomes (Cleft Development Group, 2021). The recommendation in the UK was for primary palatoplasty, secondary speech surgery to be performed within 3 months of their target threshold ages specified in the national standards, with other cleft surgical procedures categorised as priority 4 to be performed in more than 3 months (NHS England, 2018). In the USA, cleft operations have also been categorised via a tiered system with reference to the ACPA operative target timing guidelines (Zimmerman et al., 2020).

The prioritisation of surgical procedures in this study purposefully did not incorporate specific threshold timings but instead categorised procedures into high, medium and lower priorities to reflect the degree of time-sensitivity with respect to functional outcomes. There was an emphasis to prioritise multidisciplinary cleft services equally alongside surgical procedures as these complete the comprehensive approach. A common theme with both strands was a prioritisation of speech outcomes, in recognition of the crucial role globally that speech plays in the life and social functioning of children born with CL/P.

### Strengths and limitations

The main strength of this piece of work was the collaborative nature of the international working group, which was inclusive of multiple disciplines and affiliation with multiple global cleft organisations. The working group was a good size in terms of productivity, but it was not inclusive of all specialties, organisations or regions and deliberations all took place in English.

The consensus recommendations were based on common principles, but this is not an exhaustive document and therefore not a comprehensive guide to delivering cleft care protocols in LMICs during and after COVID-19. It is hoped the efforts of cleft providers in resource-constrained settings will be supported by this work to present a united and coordinated case for the provision of comprehensive cleft care to policy makers and ultimately improve safety and outcomes for patients. Ideally, there should be a focus on local protocols and guidance, therefore the relevance of these recommendations in specific environments may be limited (Truche et al., 2020).

### Further work

It is hoped that collaborative efforts such as this will galvanise the global cleft community to perform multi-centre international trials to reach a consensus on cleft care protocols and outcomes. Local outcome data collection must be encouraged to drive context-specific guidance. Finally, the efficacy of innovations highlighted by this pandemic should be explored so that they can ultimately help with the provision of global cleft care.

## CONCLUSION

The COVID-19 pandemic has had a detrimental impact on the delivery of comprehensive cleft care, which was already stretched in many areas of the world. As a global community, it is helpful for the providers of cleft care in LMICs to be able to recognise protocol adaptations that may be needed to deliver cleft care safely and elements that should be prioritised to maximise time-sensitive outcomes. A unified approach amongst global cleft care providers may help to lobby policy makers effectively at this crucial time of scarce resource allocation.

## Data Availability

All data produced in the present work are contained in the manuscript

## ACKNOWLEDGEMENTS

The authors would like to thank the Circle of Cleft professionals and the multiple organisations that support it for facilitating this work.

## SUPPLEMENTARY MATERIAL

**Supplementary Figure 1:**
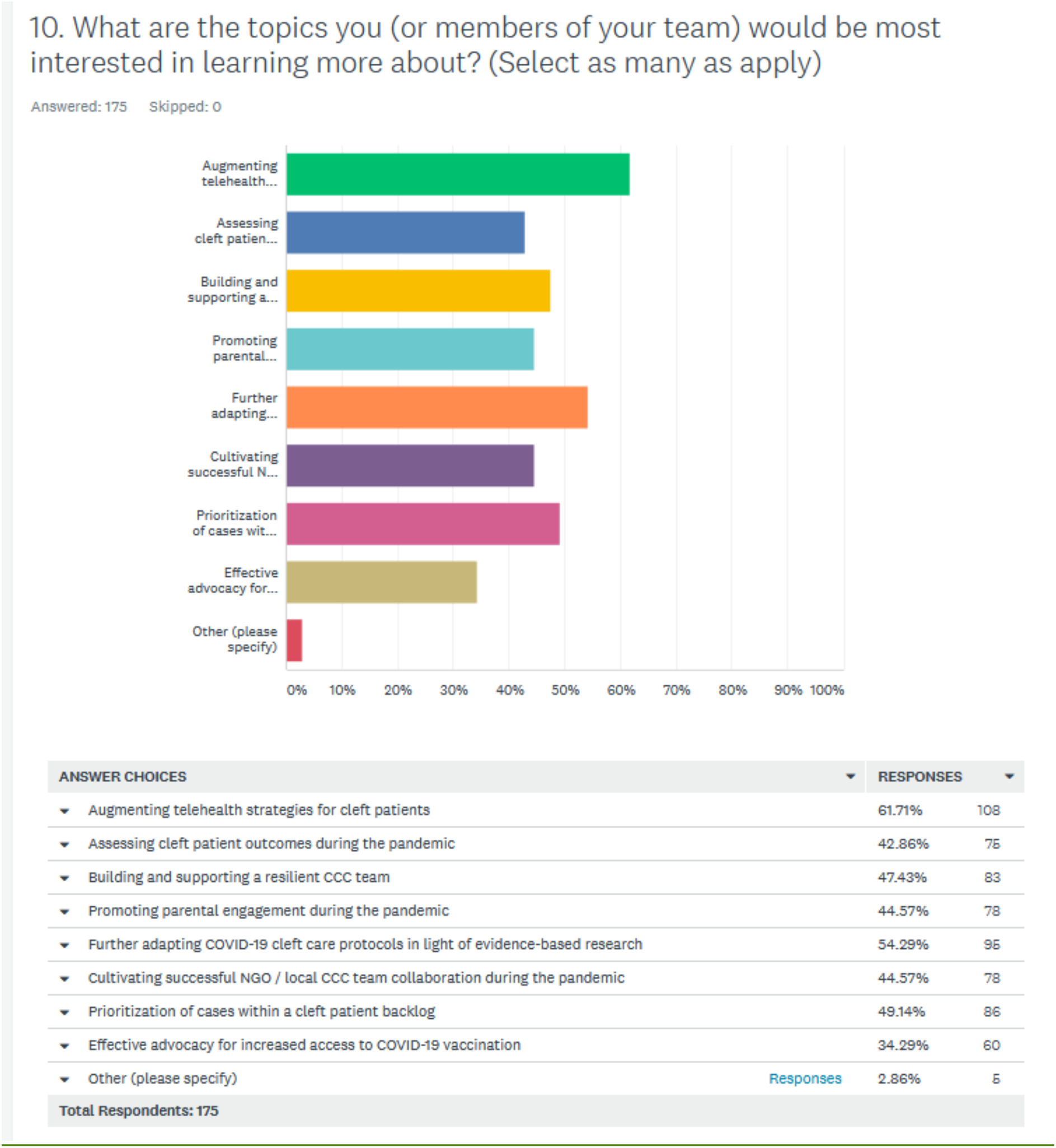
Results of Circle of Cleft Professionals COVID Survey: Question 10. What are the topics you (or members of your team) would be most interested in learning more about?

**Supplementary Figure 2:**
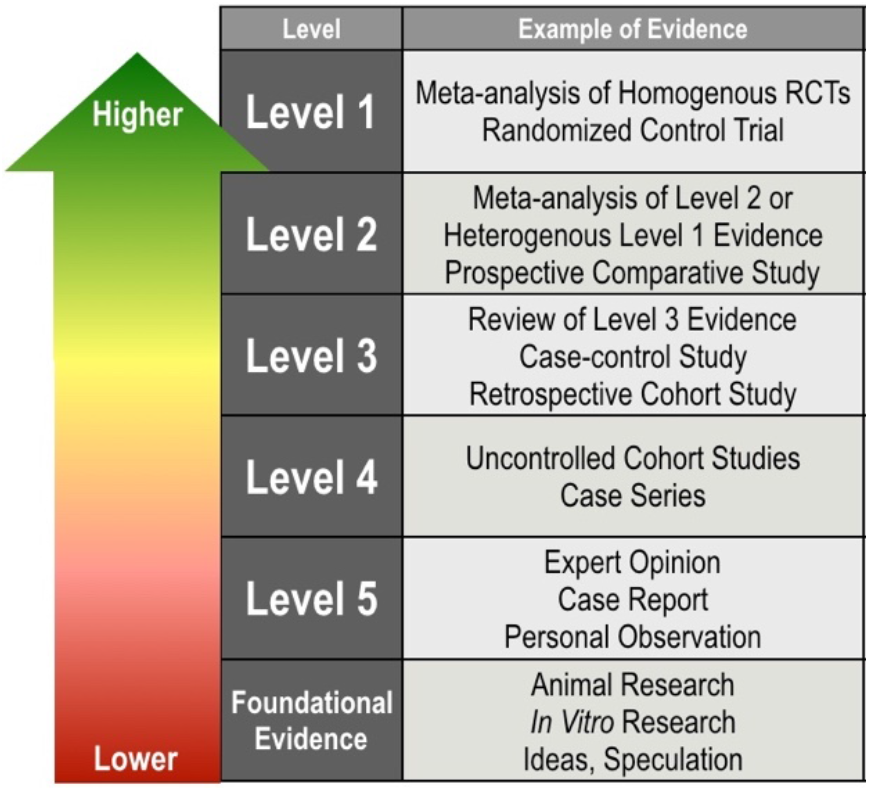
Levels of evidence used by the working group to consider literature

**Supplementary Table 1:** Evidence that timing of palatoplasty is associated with speech outcome

| Resource title | Level of Evidence | Overview of resource | Conclusion |
| --- | --- | --- | --- |
| SCANDCLEFT Trials<br>(Lohmander et al., 2017)<br>(Willadsen et al., 2017) | 1 | RCT comparing 4 2-stage surgical protocols for complete UCLP (n=<br>Hard palate closure at 12 versus 36 months was compared | No difference in resonance (nasality)<br>Later palate closure associated with poorer consonant proficiency |
| (Willadsen et al., 2018)<br>Denmark | 1 | RCT 126 children with UCLP to compare hard palate closure at 12 months versus 36 months | Early hard palate closure associated with better consonant production and fewer active cleft speech characteristic errors |
| (Willadsen, 2012)<br>Denmark | 1 | RCT 34 children with UCLP SP closure at 4 months, HP closure at 12 months or 36 months | Speech outcomes better in early repair group – 12 months. |
| (Williams et al., 2011)<br>Brazil | 1 | RCT comparing primary palate closure at 9-12 months versus 15-18 months | No statistical difference in resonance |
| (Shaffer et al., 2020) | 4 | Retrospective review 733 children born 2005-15 | Late palatoplasty associated with |
|  |  | Age at palatoplasty: Early: <11mnths, n=28;<br>Standard: 11-13mnths, n=158;<br>Late: >13mnths, n=46 | increased speech and language delay.<br>Speech sound production disorders, VPI and hearing loss were not significantly associated with age at palatoplasty |
| (Kara et al., 2020)<br>Turkey | 4 | Retrospective review of 416 cleft palate patients who had palate repair before 12 months, 12-18 months and >18 months | Palate repairs after 18 months had worse speech |
| (Swanson et al., 2017) | 4 | Retrospective review of 40 patients with submucous palate | Better speech outcomes in patients repaired before 4 years of age |
| (Follmar et al., 2015) | 4 | 13yr retrospective review of 201 internationally adopted patients 1993-2006. 183 surgery before 18 months, 18 performed after 18 months. 33% of delayed group developed VPI compared 13% of those repaired at <18 months (p=0.03) | Internationally adopted children whose palate was repaired after 18 months were more likely to develop speech symptoms of VPI. |
| (Pasick et al., 2014) | 4 | Retrospective review of 24 patients : palatoplasty at >18 months | Palate repair after 18/12 associated with significantly increased incidence of articulation errors associated with VPI |
| (Yang et al., 2013) | 4 | Retrospective review of 503 patients with non-syndromic cleft palate | Repair before 2 years of age had better velopharyngeal competence |
| (Chapman et al., 2008) | 4 | 40 children with non-syndromic cleft palate repaired primarily in younger age (mean 11 months) and older age (mean 15 months) assessed at age 3 years. | At age 3 years children who had palate surgery prior to the onset of words had better articulation and resonance outcomes than those who had surgery after the onset of words. |
| (Hardin-Jones & Jones, 2005) | 4 | 212 preschoolers with clefts | Significant relationship between age at palatal surgery and prevalence of hypernasality advocated primary repair before 13 months |
| (Rezaei et al., 2020)<br>Iran | 4 | Retrospective review of 180 patients – comparison made between primary palate repair before and after 13 months | Late surgery group had significantly worse articulation errors and more likely to have moderate/severe hypernasality |
| CRANE Database UK<br>(CRANE, 2020) | 4 | Comparing speech outcomes with timing of surgery for 789 non-syndromic children born 2007-2013. Significant difference in speech outcome with those having surgery at 14+ months doing significantly worse on all 3 UK speech standard outcomes. | Speech outcomes better in children who have speech surgery before 14 months of age. |
| (Marrinan et al., 1998) | 4 | Retrospective study of 228 non-syndromic patients with 2 surgeons. Significant linear association ($p=.025$) between age at repair and VPI. | Patients in the early repair group (8-10 months) were least likely to need secondary treatment for VPI. |
| (Dorf & Curtin, 1982) | 4 | 21 children surgery before 12 months,<br>59 surgery after 12.5 months<br>10% in early group had compensatory articulations compared to 86% in late group | Worse articulation outcomes for children whose palate surgery was completed before 12 months compared to those completed after 12 months. |
| (Peterson-Falzone, 1996) | 5 | A non-systematic review of the evidence on timing of palate surgery and speech outcome | The effects of structural constraints on phonetic and phonological development in children supports efforts towards earlier surgery (<12 months). General trend in the literature of better speech results in earlier surgery |

**Supplementary Table 2:** Evidence that the timing of primary palatoplasty is associated with maxillary growth outcome

| Study /Personal Experience | Level of Evidence | Overview | Conclusion |
| --- | --- | --- | --- |
| (Pereira et al., 2018)<br>Brazil | 1 | Single centre RCT of 64 patients to evaluate early complete palate repair (9-15 months) vs late hard palate repair (3-4 years) | Delayed hard palate repair had better dentofacial growth |
| Scandcleft Trials<br>(Küseler et al., 2019)<br>(Heliövaara et al., 2017)<br>(Karsten et al., 2017) | 1 | Part of a multi-centre RCT of 429 patients compared early (12 months) versus late (36 months) hard palate repair. Maxillary growth measured using models at 5 years and cephalometry at 8 years | No difference in maxillary growth by 8 years of age |
| (Botticelli et al., 2019)<br>Denmark but included in Scandcleft | 1 | Single centre RCT of 122 UCLP to compare hard palate closure at 12 or 36 months – assessment on models at 8 years of age | No conclusion about which was favourable – delayed repair had better transverse dimension but had a shallower morphology |
| (Salgado et al., 2019) | 4 | A systematic review of early vs delayed palatoplasty and effect on growth – 5 included observational studies<br>• Daskalogiannakis et al., 2006<br>• Holland et al., 2007<br>• Yaminishi et al., 2011<br>• Zemmann et al., 2011<br>• Bakri et al., 2014 | Conflicting results between the five studies – no conclusion made |
| (Xu et al., 2012) | 4 | Retrospective series of 46 UCLP to compare palatoplasty before and after 4 years of age | Better maxillary growth associated with later repairs |
| (Y.-F. Liao et al., 2010) | 4 | Retrospective series of 72 cUCLP to compare one stage versus two-stage | Delayed hard palate associated with |
|  |  | with delayed hard palate repair.<br>Cephalometry at 20 years. | better maxillary growth |
| (Y.-F. Liao & Mars, 2006) | 3 | A systematic review of timing of palate repair on facial growth – 15 observational studies included | No conclusive evidence that timing affects growth |
| (Y. F. Liao et al., 2006) | 4 | Retrospective series of 104 patients in Sri Lanka who had their palate repaired by 13 years of age | Later age of repair was associated with better AP growth of the maxilla |
| (Friede, 2007) | 5 | Personal perspectives of delayed hard palate closure | Advocates delayed hard palate closure at 1-1.5 years to achieve better growth |

**Supplementary Table 3:** The Federation of Surgical Specialty Associations Surgical Prioritisation System – first published June 2020. Updated in February 2021. The priority levels assigned to cleft operations are indicated both initially in July 2020 and later in February 2021.

| Priority | Time scale for surgical procedure to be performed | Initial cleft operation prioritisation July 2020 | Amendment to cleft prioritisation February 2021 |
| --- | --- | --- | --- |
| 1a | Less than 24 hours | Nil | Nil |
| 1b | Less than 72 hours | Nil | Nil |
| 2 | Less than 1 month | Nil | Primary cleft palate repair (child breaching 13 months of age),<br>Secondary speech surgery (child breaching 5 years of age) |
| 3 | Less than 3 months | Primary Palatoplasty,<br>Secondary Speech Surgery<br>Alveolar Bone Grafting | Primary Palatoplasty (child less than 12 months of age),<br>Secondary Speech Surgery (child less than 5 years of age)<br>Alveolar Bone Grafting (prior to canine eruption) |
| 4 | More than 3 months | All other cleft operations | All other cleft operations |

